# Early antibody response to SARS-CoV-2

**DOI:** 10.1101/2020.05.19.20099317

**Authors:** Ruggero Dittadi, Haleh Afshar, Paolo Carraro

## Abstract

**Background:** The role and significance of the immune response to SARS-CoV-2 infection is not yet well known.

**Methods:** We conducted a study on 46 symptomatic subjects with disease confirmed by laboratory tests, to evaluate the presence of IgG and IgM antibodies in these subjects in relation to the time elapsed since the onset of symptoms. The analytical performance of the method used in the study and the effect of two different serum and plasma matrices were also assessed.

**Results:** IgG positivity was demonstrated in 100% of cases 15 days after the onset of the disease. IgM show lower concentrations and do not exceed 77% of cases after 15 days. The analytical performance of the method used (Maglumi 800, Snibe, China) was confirmed to be good in terms of imprecision, linearity and commutability in two sample matrix.

**Conclusion:** The serological study through the search for specific IgG for SARS-CoV-2 results to be sensitive and suitable for population research and evidences that this approach can also play an important role in diagnosis.

The diagnostic performance of specific IgMs are lower.

## Introduction

The diagnosis of Covid-19 disease makes use of clinical, radiological data and molecular research of the virus on the nasopharyngeal swab, mainly with Real-Time reverse transcription Polymerase Chain Reaction (RT-PCR) (1). In the early stage of the disease, the use of serological tests has recently been proposed but its role in the diagnostic process is not yet clear. There is some indirect evidence of a protective role of specific antibodies, such as the administration of plasma immune from healed subjects to sick patients (3). This represents a very interesting challenge but definitive evidence is still lacking.

We evaluated a series of samples of subjects who presented at the Ospedale dell’Angelo in Mestre (Venice, Italy) with symptoms suggestive of SARS-CoV-2 infection, subsequently diagnosed definitively with clinical and laboratory criteria. The presence of specific IgM and IgG antibodies was assessed by a new chemiluminescence assay on MAGLUMI platform, that, from the manufacturer’s statements, detects spike and nucleocapside proteins, the main immunogens protein of coronavirus (4), that seems to correlates with neutralizing antibod responses (5). Results were related to the time elapsed since the onset of symptoms.

## Methods

Blood samples from patients admitted with probable COVID-19 from March 2 to March 29 2020 were collected in polyethylene tubes (BD Vacutainer®; Becton Dickinson, CA, USA) containing clot activator and gel separator for serum preparation. Eighteen samples were also collected in polyethylene tubes containing 3.6 mg K_2_EDTA for the comparison of blood matrices.

Patients with suggestive diagnosis and positive swab were considered with the exception of 4 cases in which in which the clinical diagnosis along with images was almost certain, despite the presence of negative RT-PCR nose-pharyngeal swab. From all these cases we selected only those with specific onset of symptoms. Therefore, 46 patients (38 males and 8 females, median age 66 years, minimum 36, maximum 89) were included, of which 30 with one blood sample, 7 with two withdrawals over the following days, 5 with 3 withdrawals, 2 with 4 withdrawals and 2 with 5 withdrawals, for a total of 77 assessed samples. Moreover, 35 healthy controls with negative swab were included (20 female, 15 males, age 24-65).

The tests were performed with a two step chemiluminescence immunoassay using magnetic microbeads coated with anti-human Immunoglobulins (IgM and IgG) and 2019-nCoV recombinant antigen labelled with luminescence substrate (MAGLUMI 2019-nCoV IgM and MAGLUMI 2019-nCoV IgG respectively). The assays were carried out on Maglumi 800 Analyzer (Snibe, Shenzen, China) according to the manufacturer’s instructions. The results were interpreted following the indications of the manufacturer: (i) IgG considered reactive if > 1.1 AU/ml; (ii) not reactive if <0.9 and (iii) doubt between 0.9 and 1.1. A cut off limit of 1.0 for IgM was proposed.

Five aliquots of three sera (S1, S2 and S3) were stored and analyzed in triplicate in 5 different analytical sessions over 7 days to verify the intra-series and total imprecision according to the CLSI EP-15-A2 protocol.

Linearity was verified by dilution test (4 dilutions 1/2 to 1/10) mixing two samples, one with concentrations of 11.8 and 4.1 AU/mL, the second of 0.12 and 0.1 AU/mL for IgM and IgG respectively.

No additional blood sampling from the care tests was performed for this study. All investigations have been conducted by following the tenets of the Declaration of Helsinki and has been complied with institutional policies.

Statistical analysis were performed with MedCalc © Software, Version 19.2.1 (MedCalc Software, Mariakerke, Belgium)

## Results

### Imprecision

*IgG:* S1 showed a mean concentration of 0.98 AU/mL an intra-assay CV= 3.8% and a total CV= 5.2%, S2 showed a mean concentration of 14.9 U/L, an intra-assay CV= 4.3% and a total CV=5.4%, S3 showed a mean concentration of 46.3 U/L, an intra-assay CV= 2.8% and a total CV=3.2%.

*IgM:* S2 showed a mean concentration of 1.67 U/L, an intra-assay CV= 5.7% and a total CV= 9.4%, S3 showed a mean concentration of 1.93 U/L, an intra-assay CV= 4.68% and a total CV=8.25%. S1 showed IgM concentrations <0.1 AU/mL.

These results confirmed the results reported by the manufacturer.

### Linearity

Dilution test showed good linearity for IgM (recovery from 95.4% to 86.1%). For IgG a deviation from linearity (recovery 74.4%) was found at concentration <0.8 AU/mL.

### Comparison EDTA vs serum

The Passing-Bablock regression showed good correlation between the two matrices suggested by the manufacturer, both for IgG (range 0.17-88.1 U/mL) and IgM (range 0.11-12.9 U/mL).

The slope of EDTA vs serum for IgG was 1.005 (95% C.I. 0.97/1.03) and the intercept of 0.0003 (95% C.I. −1.09/+0.57). The slope of EDTA vs serum for IgM was 0.95 (95% C.I. 0.91/0.99) and the intercept of 0.016 (95% C.I. −0.06/+0.06).

### Clinical correlations

None of the 35 control samples resulted positive for IgM and IgG.

Forty-seven cases (61%) were positive for IgM, while 66 cases (85.7%) were positive and 2 in gray zone for IgG.

The correlation of the results with respect to the days of the onset of the symptoms is shown in Figure 1. We have distinguished the cases of samples analyzed before 15 days of illness (Group 1) from those analyzed 15 or more days later (group 2). In Group 1 the positivity rate was 71,1% (27 samples out of 38) but we found 100% positivity rate in the remaining 39 samples collected more than 15 days after the symptoms (Group 2). The median concentration was 24.4 U/mL (25°-75° perc: 0.92-50.6) in Group 1 and 60.6 U/mL (25°-75° perc: 29.9-66.5) in Group 2 (Mann-Whitney test p=0.0003).

**Figure 1.**
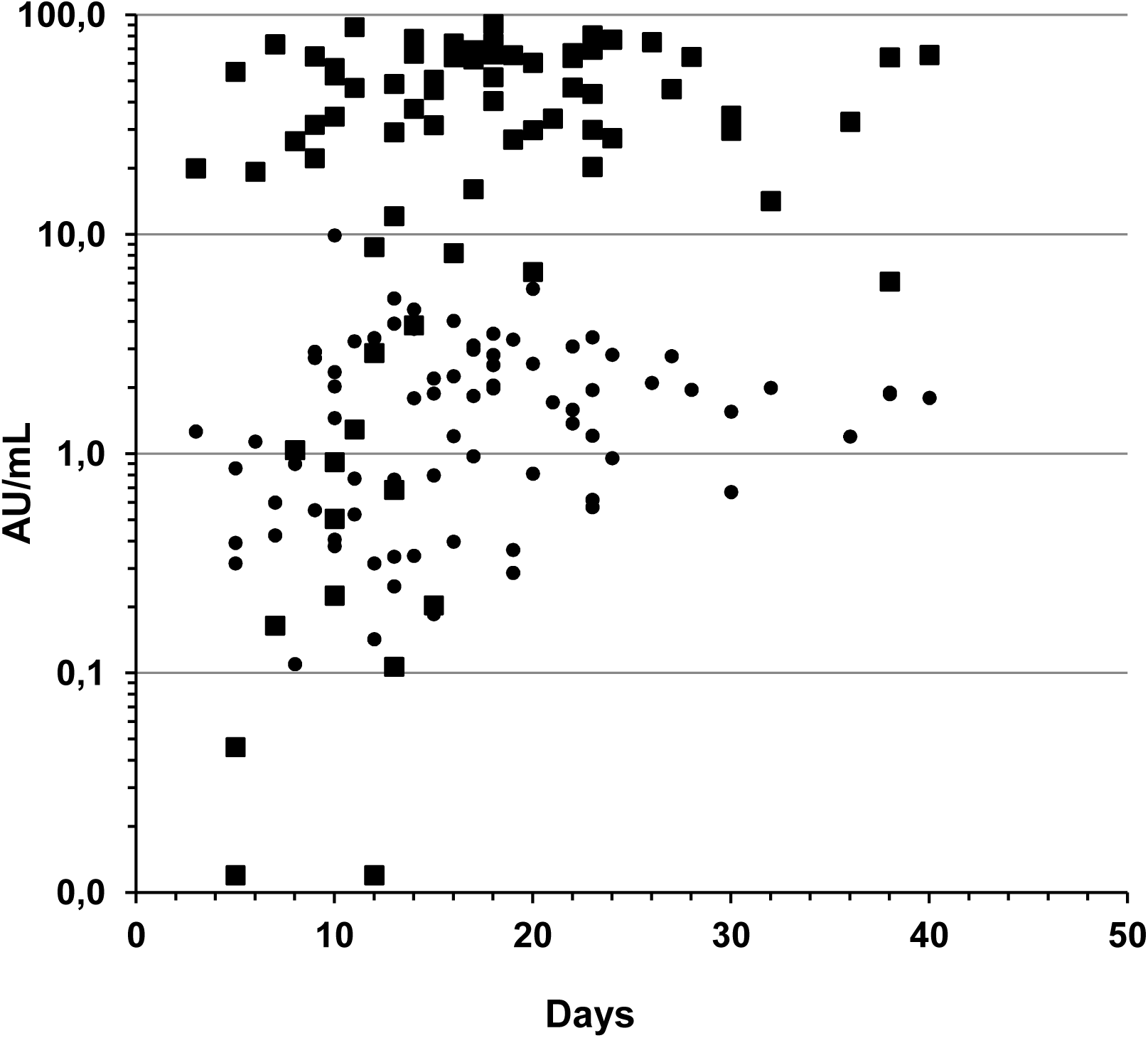
Distribution of IgM (circles) and IgG (square) in relation to the days since the onset of symptoms

Among the 10 patients with negative results, we were able to collect more than one sample for 5 patients. It is worth noting that all of them showed a rapid seroconversion.

The trend was similar for the IgM but less significant (Mann-Whytney test p=0.026). The positivity rate in Group 1 was 44.7%, with a median concentration of 0.83 U/mL (25°-75° perc: 0.39-2.36) and in Group 2 the positivity rate was 76.9%, with a median of 1.96 U/mL (25°-75° perc: 1.2-2.82).

## Discussion

The verification of the precision performances did not point out significant differences with respect to the manufacturer’s claim (overall CVs <6% within-laboratory). However, the IgM assay showed imprecision data a little higher, suggesting the possibility of identifying a gray zone between “positive” and “negative” results. Despite the limited number of cases evaluated in the control group, the absence of the detection of false positive results is encouraging. Considering COVID-19 symptomatic patients, the true positive results were 60,8% for IgM, with mainly low concentrations, and 85,1% for IgG at the first test. In particular, all the 39 samples evaluated more than 15 days after the onset of symptoms showed positive IgG results. It is moreover interesting to note that there are numerous cases with high IgG even after the first days of symptoms. It must be emphasized that the incubation time of the disease varies from 2 to a maximum of 14 days (6), and therefore it is also conceivable that cases with early high IgG may be related to patients with longer incubation times. However, in our case series the onset of the appearance of the serological response remains rather short, especially in consideration of the limited presence of IgM, which seems to have a parallel trend but with much lower concentrations than IgG. The study of 9 cases with at least 3 samples each, confirms the increase in IgG and the tendency to a plateau after 15 days (figure 2).

**Figure 2.**
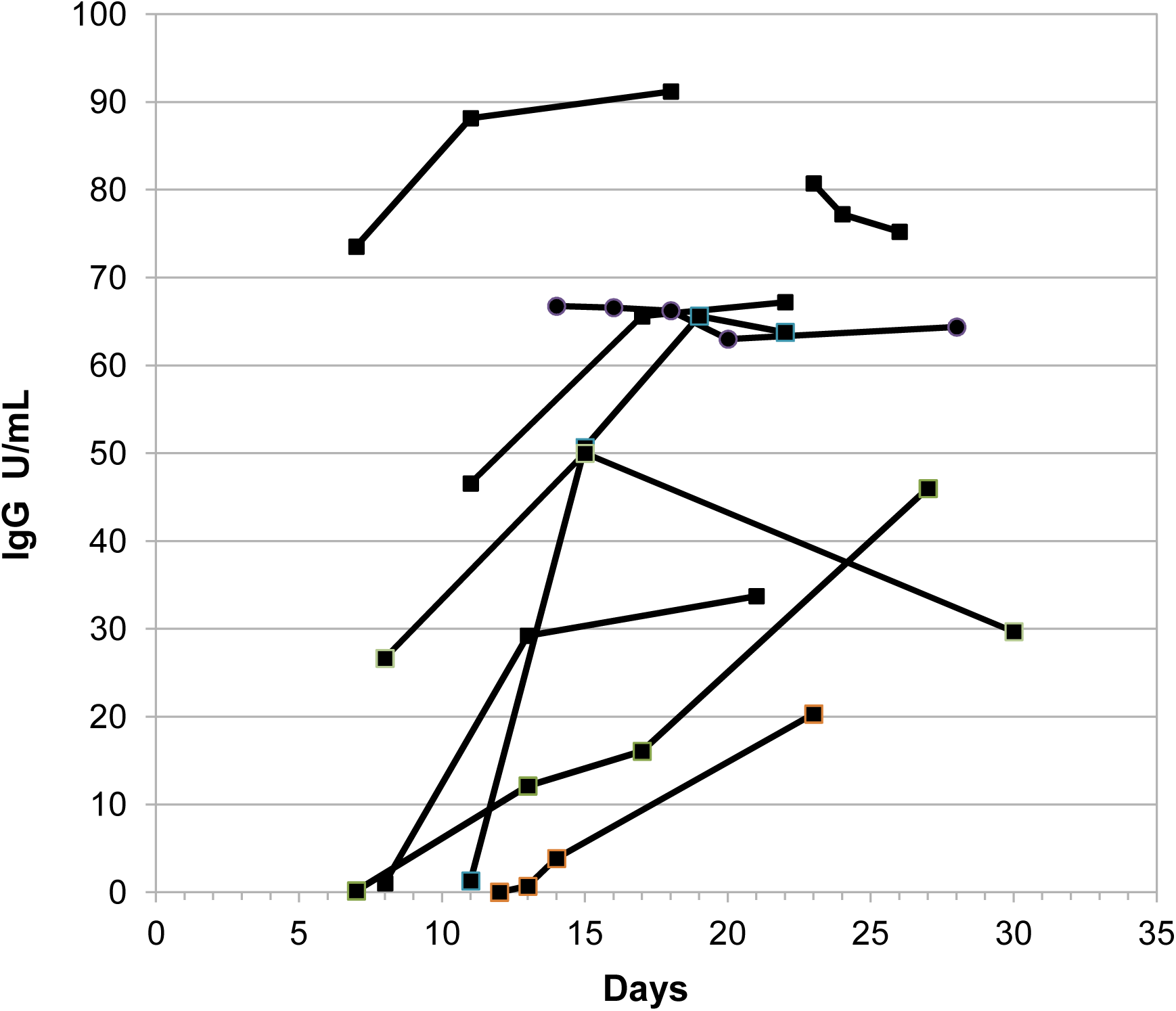
Progression of IgG values in cases with more than two samples per patients.

Recent studies are in agreement with our findings, with an almost complete seroconversion within two weeks after disease onset (7-10). In particular, in a recent report by Guo L. et al. the evidence of positive RT-PCR and the appearance of specific antibodies is very similar to that shown in our study (7). Their conclusions report a high sensitivity for the swab in the first 5.5 days of symptoms and a high sensitivity for serology in the following days. For this reason, Guo proposes the initial study of suspected patients with both diagnostic methods. Our data also seems to confirm this approach. Unlike the other studies, Padoan et al (10) accurately evaluated the method we used, and their findings were similar, both from the analytical point of view and with regard to the low expression of the IgM and the almost simultaneous appearance respect to the IgG. In fact, all the positive IgM cases except one also showed IgG positivity, so that the IgM value seems to provide a small contribution to the assessment of the immunological response in COVID-19 patients.

## Data Availability

All data of the present study are available

**List of abbreviations**
IgG: (Immunoglobulin of class G)
IgM: (Immunoglobulin of class M)
CV: (coefficient of variation)
RT-PCR: (Real-Time reverse transcription Polymerase Chain Reaction)
AU/mL: (arbitrary units per mL)
EDTA: (Ethylenediaminetetraacetic acid)
CLSI: (Clinical & Laboratory Standards Institute)

## References

1) Poston JT, Patel BK, Davis AM. Management of Critically Ill Adults With COVID-19. JAMA. 2020 Mar 26. DOI: 10.1001/jama.2020.4914

2) Zhang W, Du RH, Li B, Zheng XS, Yang XL, Hu B, Wang YY, Xiao GF, Yan B, Shi ZL, Zhou P. Molecular and serological investigation of 2019-nCoV infected patients: implication of multiple shedding routes. Emerg Microbes Infect. 2020 Feb 17;9(1):386–389.

3) C Shen, Z Wang, F Zhao. Treatment of 5 Critically Ill Patients With COVID-19 With Convalescent Plasma. JAMA doi:10.1001/jama.2020.4783. Published online March 27, 2020

4) Meyer B, Drosten C, Müller MA. Serological assays for emerging coronaviruses: challenges and pitfalls. Virus Res. 2014;194:175–83

5) Okba NMA, Müller MA, Li W et al. Severe Acute Respiratory Syndrome Coronavirus 2-Specific Antibody Responses in Coronavirus Disease 2019 Patients Emerg Infect Dis 2020; 26. doi: 10.3201/eid2607.200841

6) Lauer SA, Grantz KH, Bi Q, Jones FK, Zheng Q, Meredith HR, Azman AS, Reich NG, Lessler J. The Incubation Period of Coronavirus Disease 2019 (COVID-19) From Publicly Reported Confirmed Cases: Estimation and Application. Ann Intern Med. 2020 Mar 10. doi: 10.7326/M20-0504.

7) Guo L, Ren L, Yang S, Xiao M, Chang, Yang F, Dela Cruz CS, Wang Y, Wu C, Xiao Y, Zhang L, Han L, Dang S, Xu Y, Yang Q, Xu S, Zhu H, Xu Y, Jin Q, Sharma L, Wang L, Wang J. Profiling Early Humoral Response to Diagnose Novel Coronavirus Disease (COVID-19). Clin Infect Dis. 2020 Mar 21. pii: ciaa310. doi: 10.1093/cid/ciaa310.

8) To KK, Tsang OT, Leung WS, Tam AR, Wu TC, Lung DC et al. Temporal profiles of viral load in posterior oropharyngeal saliva samples and serum antibody responses during infection by SARS-CoV-2: an observational cohort study. Lancet Infect Dis. 2020: S1473–3099.

9) Loeffelholz MJ, Tang YW. Laboratory diagnosis of emerging human coronavirus infections – the state of the art. Emerg Microbes Infect. 2020;9:747–756

10) Padoan A, Cosma C, Sciacovelli L, Faggian D, Plebani M. Analytical performances of a chemiluminescence immunoassay for SARS-CoV-2 IgM/IgG and antibody kinetics.. Clin Chem Lab Med. 2020 Apr 16. pii: /j/cclm.ahead-of-print

